# Retina-arrestin is not a CD8+ T-cell autoantigen in HLA-A29-positive birdshot chorioretinitis

**DOI:** 10.1101/2022.10.21.22281266

**Authors:** W.J. Venema, S. Hiddingh, G.M.C. Janssen, J. Ossewaarde, N. Dam van Loon, J.H. de Boer, P.A. van Veelen, J.J.W. Kuiper

## Abstract

**Background:** HLA-A29-positive *birdshot chorioretinitis* (BCR) is an inflammatory eye disorder that is generally assumed to be caused by an autoimmune response to HLA-A29-presented peptides from retinal arrestin (SAG), yet the epitopes recognized by CD8+ T cells from patients remain to be identified.

**Objectives:** The identification of natural ligands of SAG presented by HLA-A29. To quantify CD8+ T cells reactive to antigenic SAG peptides presented by HLA-A29 in patients and controls.

**Methods:** We performed mass-spectrometry based immunopeptidomics of HLA-A29 of antigen-presenting cell lines from patients engineered to express SAG. MHC-I Dextramer technology was utilised to identify antigen-specific CD8+ T cells reactive to SAG peptides in complex with HLA-A29 in a cohort of BCR patients, HLA-A29-positive controls, and HLA-A29-negative controls.

**Results:** We report on the naturally presented antigenic SAG peptides identified by sequencing the HLA-A29 immunopeptidome of antigen-presenting cells of patients. We show that the N-terminally extended SAG peptide precursors can be trimmed *in vitro* by the antigen-processing aminopeptidases ERAP1 and ERAP2. Unexpectedly, no antigen engagement by CD8+ T cells upon stimulation with SAG peptides was observed in patients or HLA-A29-positive controls. Multiplexed HLA-A29-peptide dextramer profiling of a case-control cohort revealed that CD8+ T cells specific for these SAG peptides were neither detectable in peripheral blood nor in eye biopsies of patients.

**Conclusions:** Collectively, these findings demonstrate that SAG is not a CD8+ T cell autoantigen and sharply contrast the paradigm in the pathogenesis of BCR. Therefore, the mechanism by which HLA-A29 is associated with BCR does not involve SAG.

## Introduction

Birdshot chorioretinitis (BCR) is a rare inflammatory eye disease characterized by progressive T-cell infiltrates in the posterior eye segment, destruction of photoreceptors, gradual deterioration of retinal function, and declining visual abilities ^1–3^. Selective inhibition of T cells by Cyclosporin A is effective for treatment of birdshot chorioretinitis which supports a T-cell mediated autoimmune aetiology ^4,5^.

The most distinguished molecular trait of BCR is its extreme genetic association with the *Human Leukocyte Antigen class I A*29 allele* (HLA-A29). This gene-disease link is so robust that testing positive for HLA-A29 is considered essential for diagnosis ^2,6^. Other more recently identified susceptibility genes are the closely related *endoplasmic reticulum aminopeptidase* (*ERAP)1* and *ERAP2* genes ^7–9^, which encode enzymes specialised to shorten intracellular precursor peptides to a length that facilitates or prevents their binding to HLA class I molecules. Collectively, these molecular features strongly implicate CD8+ T-cell activation by self-antigen presentation via HLA-A29 as a key disease mechanism for BU. To date, however, the evidence for pathogenic CD8+ T cell involvement in BCR remains limited to the detection of CD8+ T-cell infiltrates in eye tissues of BCR patients ^10–12^.

The identification of bona fide CD8+ T cell autoantigens is challenging and highly labour and resource intensive. Consequently, to date, very few autoantigens for MHC-I-associated conditions are identified ^13–15^. A long presumed (since 1982) autoantigen for BCR is the *retinal arrestin* (also known as the *retinal Soluble (S-)antigen* (SAG)), which is abundantly and specifically expressed by photoreceptors. Immunisation with SAG in primates or rodents causes *experimental autoimmune uveitis* characterised by T-cell immunity directed towards the retina, with close resemblance to BCR ^16,17^. Pharmacological inhibition of T cells blocks SAG-induced experimental uveitis ^17^. Peripheral blood lymphocyte cultures from BCR patients show strong proliferation in response to stimulation with SAG ^18^ and *in vitro* binding studies support that synthetic peptides identical to fragments of the SAG can bind to HLA-A29 ^19^. Consequently, SAG is generally viewed as a causal autoantigen for BCR ^2,18–20^, but there are no studies have provided evidence for SAG peptide recognition by CD8+ T cells in the context of HLA-A29.

In this study, we characterised the self-antigen-presenting function of HLA-A29 by lentiviral transduction of full-length SAG in antigen presenting cells from patients. We used mass-spectrometry based peptide sequencing of ligands eluted from HLA-A29 at the cell surface for unbiased SAG ligand discovery. We report on the identification of two HLA-A29-presented peptides from SAG and reveal that these peptides in complex with HLA-A29 are not recognized by CD8+ T-cells of patients with BU.

## Materials & Methods

### Patient-derived antigen presenting cell lines

This study was performed in compliance with the guidelines of the Declaration of Helsinki and has the approval of the local Institutional Review Board (University Medical Center Utrecht). Antigen presenting cell lines were generated by the induction of EBV-immortalised lymphoblastoid cells (LCLs) with B95-8 marmoset-derived EBV supernatant as described earlier ^21^. Briefly, peripheral blood mononuclear cells (PBMCs) from two BCR patients were cultured in B95-8 marmoset-derived EBV supernatant with *Roswell Park Memorial Institute 1640* medium (RPMI 1640) supplemented with 10% heat-inactivated fetal bovine serum (FBS, Biowest Riverside), and 1µg/ml cyclosporine to remove T cells. LCL cluster formation was monitored by light microscopy and established patient-derived LCLs maintained in RPMI 1640 (10% FBS + 1% penicillin/streptomycin (Thermo Fisher Scientific)). Genotype data for single nucleotide polymorphisms at *5q15* (**Table 1**) from the two patients were obtained from a previous genotyping study ^8^. High-resolution HLA class I typing (**Table 1**) was performed on genomic DNA of cell lines using SSO DNA typing (LABType HD; One Lambda).

**Table 1.**
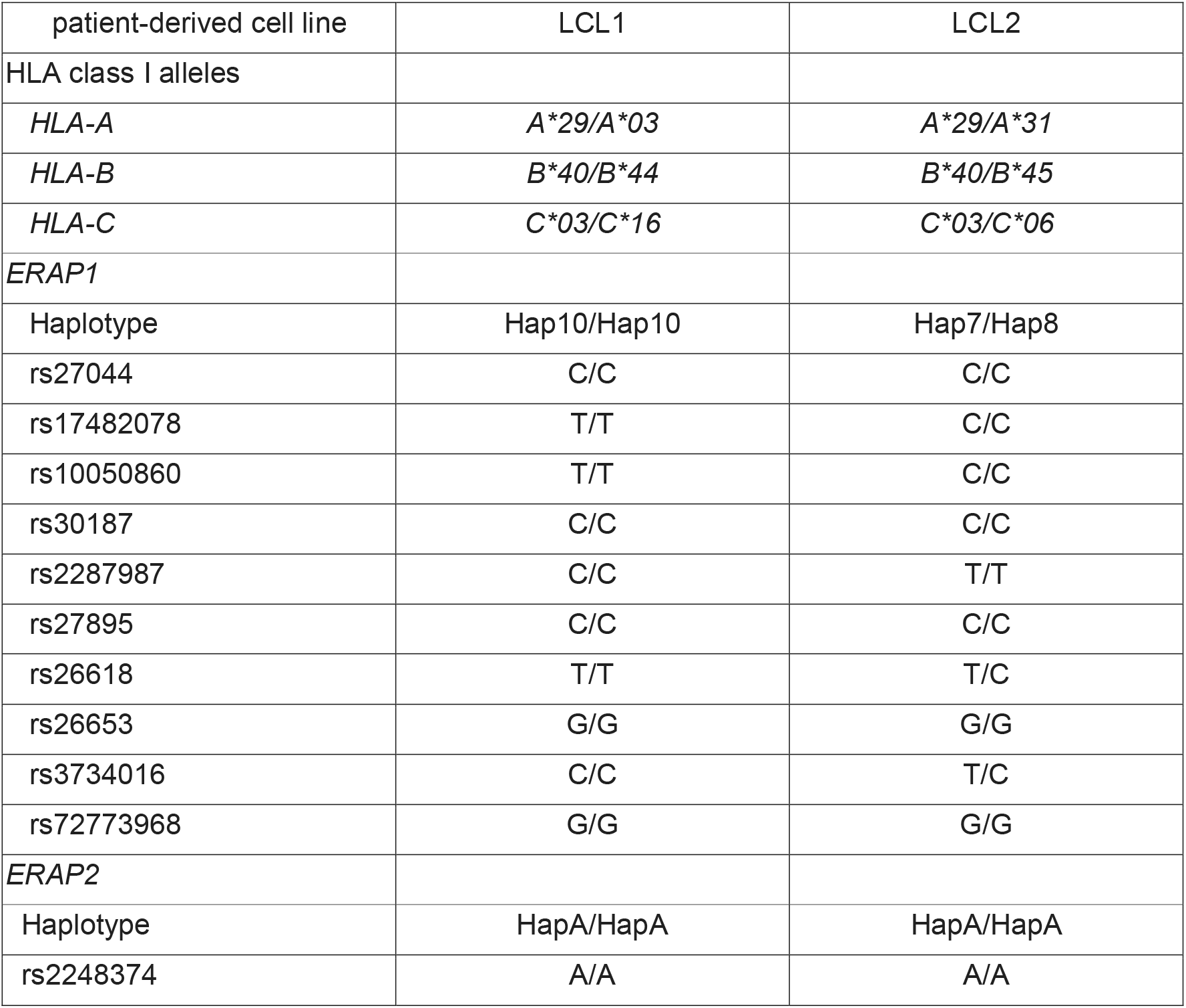
Demographics and genotype data of the patient-derived lymphoblastoid cell lines used for HLA-A29 immunopeptidomics in this study. The genotype data for HLA class I alleles and key polymorphisms (and derived haplotypes) are indicated.

| patient-derived cell line | LCL1 | LCL2 |
| --- | --- | --- |
| HLA class I alleles |  |  |
| *HLA-A* | *A\*29/A\*03* | *A\*29/A\*31* |
| *HLA-B* | *B\*40/B\*44* | *B\*40/B\*45* |
| *HLA-C* | *C\*03/C\*16* | *C\*03/C\*06* |
| *ERAP1* |  |  |
| Haplotype | Hap10/Hap10 | Hap7/Hap8 |
| rs27044 | C/C | C/C |
| rs17482078 | T/T | C/C |
| rs10050860 | T/T | C/C |
| rs30187 | C/C | C/C |
| rs2287987 | C/C | T/T |
| rs27895 | C/C | C/C |
| rs26618 | T/T | T/C |
| rs26653 | G/G | G/G |
| rs3734016 | C/C | T/C |
| rs72773968 | G/G | G/G |
| *ERAP2* |  |  |
| Haplotype | HapA/HapA | HapA/HapA |
| rs2248374 | A/A | A/A |

### S-antigen lentiviral transduction

We seeded HEK-293T cells into 10 cm dishes at a concentration of 2×10^6^ cells per dish overnight in *Dulbecco’s Modified Eagle Medium* (DMEM, Thermo Fisher Scientific). Cells were transfected in serum-free DMEM using Lipofectamine 2000 (Thermo Fisher Scientific) with 2nd generation packaging vectors (8.33 µg psPAX2 packaging vector and 2.77 µg pMD2.G envelope vector at a ratio of 4:1) and 2 µg of the transfer vector (Lenti ORF clone of Human S-antigen mGFP tagged, RC220057L2, *Origene*) and cultured at 37°C, 5% CO_2_ for 24 hours. We replaced the medium with 10 mL DMEM supplemented with 10% FBS and collected the conditioned medium containing lentiviral particles 48 hours after transfection. We added an additional 10 mL of fresh culture medium, which was harvested after 12 hours. The first and second rounds of harvested supernatants were combined and cleared by centrifugation at 1500 rpm for 5 minutes at 4°C. Cleared supernatant was passed through a 0.45 μm filter and concentrated by ultracentrifugation (Beckman Coulter Optima centrifuge, SW32Ti rotor) for 120 minutes at 32,000□rpm. After removal of supernatant, virus pellets were resuspended in 1 mL RPMI (10% FBS, 1% penicillin/streptomycin) and stored until use at −80°C. Established LCLs (1×10^6^ cells) were seeded in a 24-well plate and transduced with lentiviral particles and polybrene (final concentration 6 µg/mL). After 24 hours of culture, the medium was replaced for fresh RPMI (10% FBS, 1% penicillin/streptomycin) and the cells were cultured for another 3 days, without exceeding a cell concentration of 1.5×10^6^ cells/mL. Successful lentiviral transduction was evaluated by detection of GFP-positive cells by fluorescent light microscopy. We sorted GFP-positive LCLs using the BD FACSAria™ III sorter.

### Western blot analysis

Immunoblotting was performed to assess protein levels of S-antigen after lentiviral transduction (S-antigen fusion protein with GFP), ERAP2 and □-tubulin (loading control) with the following antibodies: ERAP2 and α-mGFP to detect the fusion protein S-antigen-GFP. Cell lysates of LCL1 and LCL2 were prepared using the NP40 lysis buffer (1% NP40, 135 mM NaCl, 5 mM EDTA, 20 mM Tris-HCl, pH=7.4) complemented with 1:25 cOmplete protease inhibitor cocktail (Roche). The protein lysates (10 µg/lane) were separated on a 4%–20% Mini-PROTEAN TGX gel (Bio-Rad Laboratories) and transferred to a polyvinylidene difluoride membrane (Immobilon-P PVDF, Millipore). Precision Plus Protein All Blue Standards was used as the protein marker (Bio-Rad Laboratories). Membranes were blocked in 5% nonfat dry milk in TBS-T and incubated overnight at 4°C with antibodies targeting ERAP2 (1:2500, AF3830, R&D Systems), □-mGFP (1:2000, clone 2F6, TA180076, Origene). Detection of ERAP2 was done using anti-goat secondary antibody staining conjugated to horseradish peroxidase (HRP) (1:5000, DAKO) and □-mGFP was measured with anti-mouse secondary antibody conjugated to HRP (1:5000, DAKO). Protein bands were detected with Amersham ECL™ Prime Western Blotting Detection System (RPN2236, GE Healthcare) on the ChemiDoc Gel Imaging System (Bio-Rad Laboratories).

### HLA-A29 immunopeptidome analysis

The immunopeptidome analysis of HLA-A29 was performed as described previously ^21^. Briefly, LCL1-SAG+ cells were cultured in a medium supplemented with stable isotope labelled amino acids and combined with unlabelled CRISPR-edited cells for SILAC-labelled immuno peptidomics as described in detail in *Venema et al*. ^21^. LCL2-SAG+ cells were cultured in unlabelled conditions. Frozen cell pellets (−20°C) of 1-2 × 10^9^ SAG-positive LCLs were lysed in Tris-HCL buffer (Tris-HCl 50 mM, NaCl 150 mM, EDTA 5mM) with 0.5% Zwittergent® 3-12 Detergent and Roche cOmplete Protease Inhibitor Cocktail and 4°C for 2 hours. Lysates were cleared by centrifugation (10 min. 2,500 rpm, 4°C). Supernatant was harvested and centrifuged for 40 min (30,000 x g, 4°C) and pre-cleared with a 2mL CL4B column. The final supernatant was subjected to the HLA-A29-binding mAb *DK1G8* ^21,22^ (IgG1) immunoaffinity column for standard immunoaffinity purification ^23^. Columns were washed and HLA-A29-peptide complexes eluted from the column. We dissociated complexes with 10% acetic acid filtered for peptides using a 10 kDa membrane (Microcon YM-10). Freeze dried filtrate was dissolved in 50 mM ammonium bicarbonate (pH 8.3) and subjected to C18 reverse phase columns (Oasis HLB, Waters, Milford, MA). Peptides were eluted from the C18 columns using 400 μl 10/90/0. water/acetonitrile/formic acid, v/v/v., 20/80/0.1 and 50/50/0.1 water/acetonitrile/formic acid, v/v/v, lyophilized and finally dissolved in 95/3/0.1 v/v/v water/acetonitrile/formic acid.

Mass spectrometry was performed using on-line C18 nanoHPLC MS/MS with a system consisting of an Easy nLC 1200 gradient HPLC system (Thermo, Bremen, Germany), and a (for LCL1) LUMOS mass spectrometer (Thermo) or (for LCL2) a Q Exactive Plus Hybrid Quadrupole-Orbitrap as described in detail in Venema *et al*. ^21^.

### Mass Spectrometry Data Analysis

Processed mass spectrometry (MS) data from LCL1 (available via https://github.com/jonaskuiper/ERAP2_HLA-A29_peptidome) were obtained by filtering peptides detected in both biological replicates for ‘heavy’ labelled amino acids (see **Supplemental Figure 1**). Peptide data from LCL1 and LCL2 were filtered for peptides with a Best Mascot ion score >35 and length between 8-11 amino acids (typical length for T-cell epitopes)^24,25^ (**Supplemental Table 1 and 2**).

**Table 2.**
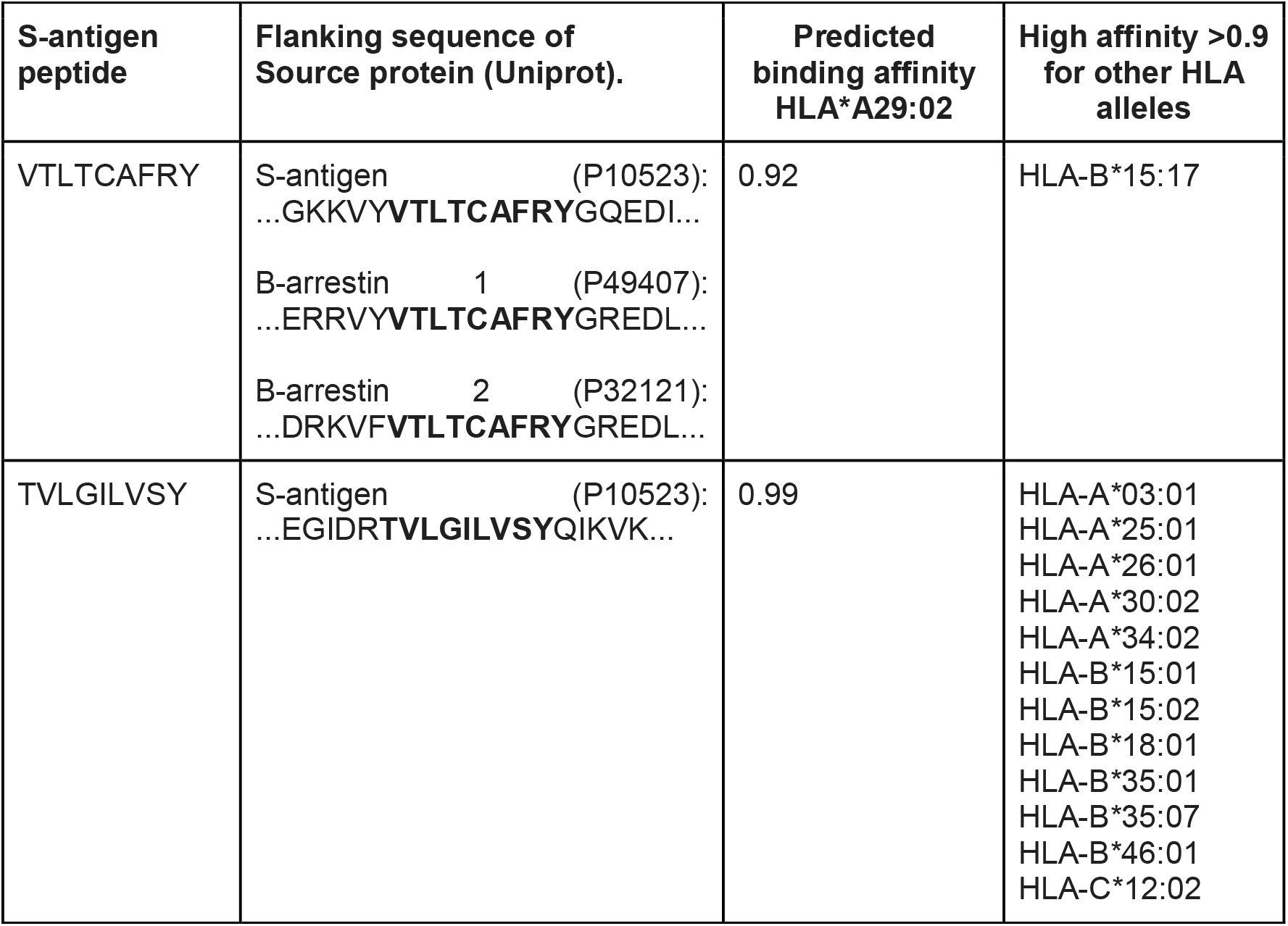
Identified S-antigen derived peptides in immunopeptidome data of LCL1 and LCL2. The flanking sequence in the source protein(s) is shown. The predicted binding affinity [range 0-1, from HLAthena^26^) for HLA*A29:02 and other class I alleles are also provided.

### ERAP1 and ERAP2 S-antigen peptide digestions

Recombinant ERAP1 (2334-ZN-010, R&D Systems) and ERAP2 (3830-ZN-010, R&D Systems) were used for digestion of SAG peptides (VTLTCAFRY and TVLGILVSY) and their respective natural N-terminal extended 10-mer precursor (all custom order from Pepscan Presto, NL) (**Key Resources Table**). We used the peptide SVLKSLPFTII and RGRFSGLLGR as positive controls for ERAP1 and ERAP2, and SVLKSLPFTII was also used to evaluate potential tryptic activity. Each peptide (1 µg) was individually digested with either ERAP1, ERAP2 or a combination of both ERAPs (100 ng per condition). Reactions were terminated by addition of 5% trifluoroacetic acid (TFA) after 60 minutes incubation at 37°C. N-terminal peptide trimming was assessed using mass spectrometry comparing peptide cleavage between T0 and T1. All reactions have been performed *in duplo* in separate experiments.

### HLA-A29-peptide dextramer analysis

To determine the presence of antigen-specific T cells reactive to retinal S-antigen peptides, peripheral blood mononuclear cells (PBMCs) from Birdshot patients, anonymous HLA-A29-positive controls (Buffy coat, Sanquin, Amsterdam), and HLA-A29-negative anonymous healthy blood donors with no history of ocular inflammatory disease served as controls (HC) were isolated from heparinized venous blood by standard ficoll gradient centrifugation immediately after blood withdrawal and stored in liquid nitrogen. For flow cytometry analysis the liquid nitrogen stored PBMCs were quickly thawed and washed once with ice cold phosphate buffered saline. The cells were stained with Fixable Viability Dye eFluor 450 (eBioscience) to determine the viability. After 10 minutes of incubation, samples were stained with APC-labeled dextramer *HLA-A*29:02-TVLGILVSY*, PE-labeled dextramer *HLA-A*29:02-VTLTCAFRY* and a FITC-labelled negative control (*HLA-B*08:01* dextramer loaded with nonsense AAKGRGAAL peptide) (Immudex, DK). The incubation was performed in FACS Buffer (1% bovine serum albumin (BSA) and 0.1% sodium azide in PBS) at room temperature for 10 minutes, followed by washing and further surface staining with V500-labeled anti-human CD8 (Becton Dickinson), PerCP-Cy5.5-labeled anti-human CD4 (BioLegend), BV605-labeled anti-human CD3 (BioLegend), PE-CF594-labeled anti-human CD56 (Becton Dickinson) and a dump channel of APC-eF780-labeled antibodies (CD14 and C19; eBioscience) for 20 minutes at 4°C. Flow cytometric analyses were performed on the BD FACSAria™ III and the data were analyzed using FlowJo software (TreeStar Inc.). Fluorescence minus one (FMO) control samples were used to set the dextramer gates.

As a control, CompBead Plus Anti-Mouse Ig,κ beads and unlabelled negative control beads (both BD Biosciences) were mixed with anti-human HLA-A,B,C antibody (clone W6/32; BioLegend), followed by washing and further staining with either one of the dextramers. Analysis of the beads was performed on the BD FACSAria™ III. For the analysis of aqueous humour (n=1), and vitreous fluid (n=1), samples from two BCR patients were stained and analysed as described above. However, only one wash step after staining was performed to minimise cell loss given the limited number of cells present in the samples.

### PBMC stimulation with S-antigen peptides

PBMCs from 12 BCR patients and 6 HLA-A29-positive controls were seeded in 96-well flat-bottom plates (1×10^6^ cells/well) and cultured in RPMI 1640 medium supplemented with 5 IU/ml IL-2 and 10% human AB serum. The cells were stimulated with either one of the synthetic peptides TVLGILVSY or VTLTCAFRY (Pepscan Presto, NL) for 48 hours at a concentration of 10 ng/ml. After 48 hours of peptide stimulation, cells were re-stimulated for 4 hours with 20ng/mL ionomycin and 1µg /mL phorbol myristate acetate (PMA) (both from Sigma Aldrich). GolgiStop (BD Biosciences; 2µL in 150µL culture medium) was added after 30 minutes and the cells were cultured for an additional 3.5 hours before the intracellular cytokine staining was performed.

Cells were surface stained with APC/Cy7-labeled anti-human CD8 (Becton Dickinson), BV785-labeled anti-human CD4 (BioLegend), BV605-labeled anti-human CD3 (BioLegend), fixed, and permeabilized in fixation buffer (Becton Dickinson) and intracellularly stained using antibodies against TNFα (BioLegend), IL-17A (eBioscience) and IFNγ (Becton Dickinson). Flow cytometric analyses were performed on the BD LSR Fortessa™ Cell analyzer (BD Bioscience). Data were analyzed using FlowJo software (TreeStar Inc.).

### Peptide prediction algorithms and Statistical analysis

Peptide binding to HLA alleles was predicted using neural-network based prediction algorithm HLAthena ^26^. HLAthena provides a binding score metric ‘MSi’ between 0-1 with MSi>0.9 considered strong binding. Prediction of antigen processing of N-terminally extended precursor peptides was conducted by subjecting the full amino acid sequence of of SAG (Uniprot: P10523, ARRS_HUMAN), □-arrestin-1 (Uniprot: P49407, ARRB1_HUMAN) and □-arrestin-2 (Uniprot: P32121, ARRB2_HUMAN) to NetChop - 3.1 web server ^27^ using the *Cterm* method, which best predicts cellular antigen processing outcome (proteasome+proteases) ^28^. The mean protein copy number of beta-arrestin 1 and beta-arrestin 2 isoforms in purified peripheral blood immune cell subsets were calculated using the mass-spectrometry-based proteomic data as reported by Rieckmann and co-workers ^29^. Group differences were assessed using the Kruskal-Wallis rank sum test with the *kruskal*.*test()* function in base R or a Dunn’s Kruskal-Wallis Multiple Comparisons by *dunnTest()* function in the *FSA R* package^30^. *P* values were adjusted using the bonferroni method.

### Data availability

Descriptions of how to reproduce the analysis workflows (showing code and R package version numbers) using data underlying the figures presented in this paper are available at *DataverseNL* via: *soon available*.

## Results

### Peptides from the retinal S-antigen (SAG) are bona fide ligands of HLA-A29

We reasoned that a powerful way to discover peptide antigen ligands derived from the retinal S-antigen (SAG) would be to determine if they can be presented by HLA-A29 using patient-derived antigen presenting cells (**Fig. 1A**). To do so, we initiated the study by using immunopeptidome data from an *HLA-A29:02*-positive birdshot chorioretinitis patient derived lymphoblastoid cells (LCL1, female, in their 80s) that we subjected to lentiviral transduction for stable expression of full-length SAG protein (SAG-GFP fusion protein) (**Fig. 1B,1C**). We filtered the amino peptidome data for 8-11 amino acids long peptides (i.e., typical length for T-cell epitopes)^25^ with an ion Mascot Score >35 (n=820 unique peptide ligands)(**Fig. 2A** and **Supplemental Figure 1**) and identified the 9-mer peptides VTLTCAFRY [amino acid positions 63-71 in SAG] and TVLGILVSY [323-331] that matched with the amino acid sequence of SAG (**Table 1, Supplemental Table 1**).

**Figure 1:**
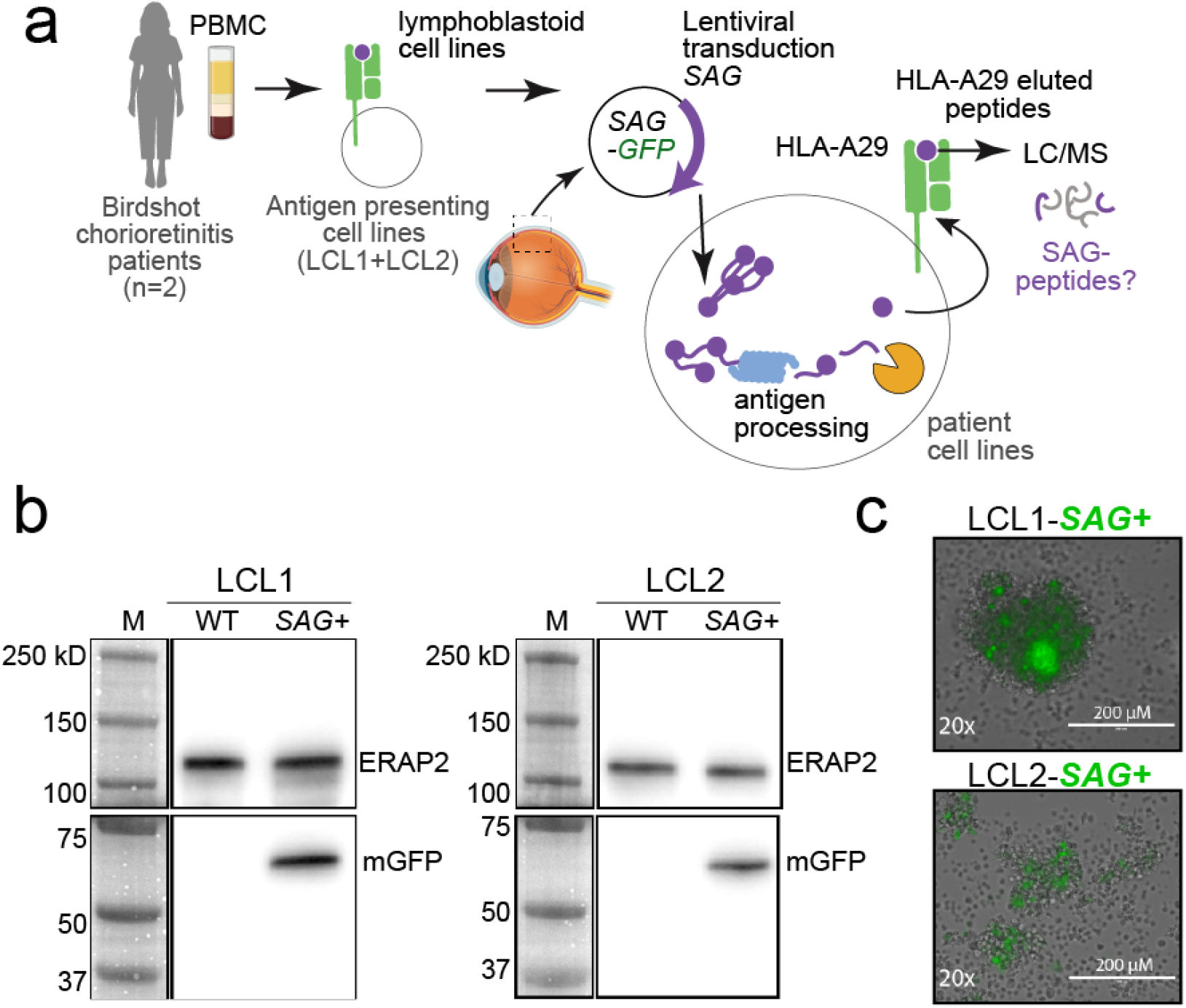
Study design and generation of patient-derived antigen presentation models for SAG. **A**) Schematic overview of the generation of a birdshot chorioretinitis patient-derived lymphoblastoid cells [LCL1 and LCL2] expressing the retinal S-antigen (SAG)-GFP fusion protein by lentiviral transduction. We used mass-spectrometry-based peptide sequencing (i.e., *immuno-peptidomics*) of HLA-A29-eluted peptides for identification of SAG-derived ligands of HLA-A29. **B**) Western blot of SAG+mGFP fusion protein (27.3 kD mGFP + 44.6 kD S-antigen) expression and ERAP2 (control) in “WT” and LCL1s transduced with SAG. Two separate blots using the same protein lysates for each sample were run in parallel for the detection of either anti-ERAP2 or anti-mGFP. **C**) Brightfield microscope image with GFP overlay of LCLs after stable expression of SAG-GFP.

**Figure 2.**
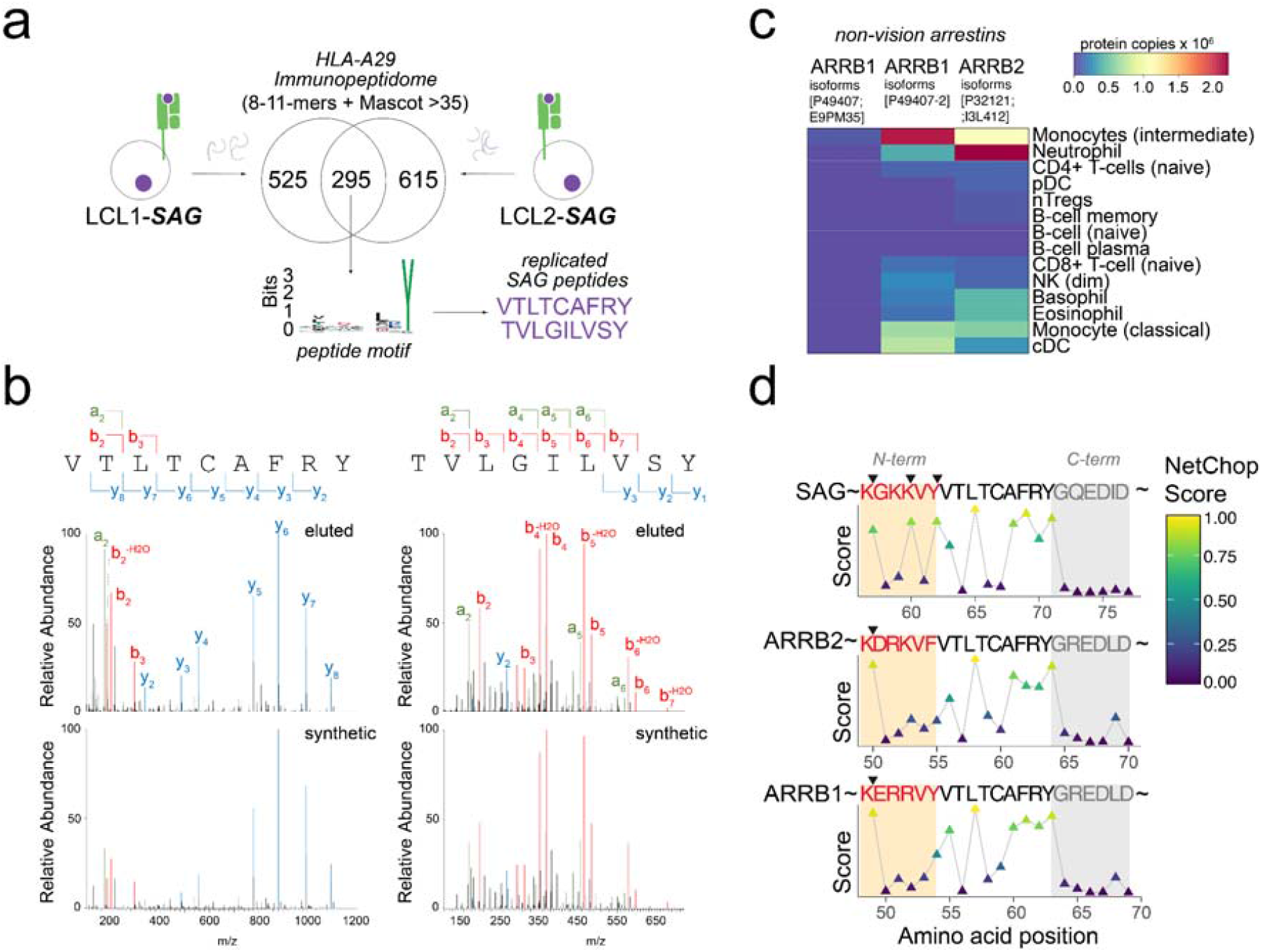
Identification of two SAG derived ligands of HLA-A29. **A**) Venn diagram of HLA-A29 immunopeptidomes (8-11 mers with Mascot Ion score >35) of LCL1-*SAG* and LCL2-*SAG*. Detailed filtering steps for peptides in the immunopeptidome data is provided in **Supplemental Figure 1**. A sequence logo is shown based on the non-redundant list of 9-mers detected in both cell lines and includes the SAG-derived peptides (VTLTCAFRY, TVLGILVSY) highlighted in purple. **B**) The MS/MS spectrum of the peptide fragment ions derived from the SAG peptides identified in elutions from HLA-A29 immunoprecipitations of the patient derived cell lines (“eluted”) and from recombinant peptides with identical amino acid sequence (“synthetic”). The relative abundance and mass/charge (m/z) of characteristic (a-, b-, and y-) ions from the fragmentation of the peptides are highlighted. Detailed MS/MS fragment spectrum is shown in **Supplemental Figure 2. C**) Heatmap showing the the mean protein copy numbers for protein isoforms of □-arrestin-1 and -2 (ARRB1, -2) in peripheral blood immune subsets (data from Rieckmann *et al*.^29^ **D**) The predicted in vivo whole-cell proteolysis (proteasomal + all other proteases) of SAG, and □-arrestin-1 and -2 (ARRB1, -2) of full-length protein by *NetChop Cterm* algorithm^27^. The predicted cleavage sites in the source protein N-terminally of the identified ligand VTLTCAFRY that exceed the threshold (>0.5) are highlighted with black triangles.

To validate these findings, we generated LCLs from a second *HLA-A*29:02*-positive birdshot chorioretinitis patient (LCL2; female, in their 60s) (**Table 1**) and SAG was also stably expressed by lentiviral transduction in this cell line (**Fig. 1B and 1C**). Using mass spectrometry analysis, we sequenced peptides eluted from anti-HLA-A29 immunoprecipitations from LCL2 and filtered the immunopeptidome data (n=910 unique 8-11-mers, **Supplemental Figure 1)** and detected 295 peptides (Ion Mascot Score >35) in common with LCL1 (**Supplemental Table 2** and **3**). This independent analysis also detected VTLTCAFRY (SAG[63-71]) and TVLGILVSY (SAG[323-331]) as the only SAG derived peptides in the HLA-A29 immunopeptidome data of LCL2. Comparing the tandem mass spectrometry (MS/MS) spectra of the SAG ligands identified in the elutions from HLA-A29 immunoprecipitations with the fragment-spectrum we generated from their synthetic peptide analogues supported accurate identification of SAG derived peptides bound to HLA-A29 (**Fig. 2B, Supplemental Figure 2**). In silico binding analysis by *HLAthena* ^26^ corroborated that these peptides were also predicted as very strong ligands for HLA-A29 (**Table 2**).

Using the amino acid sequence of these peptides as a query in BLAST^31^ revealed that TVLGILVSY is unique to SAG. In contrast, the peptide sequence of VTLTCAFRY is also present in the amino acid sequence of other non-vision arrestins (*e*.*g*., □-arrestin-1 and -2).

Although we are formally unable to assign VTLTCAFRY to the SAG, □-arrestins are low expressed in B cells (**Fig. 2C**) and this peptide has not been reported in HLA-A29 immunopeptidome studies of lymphoblastoid cell lines ^26,32–34^. Because the amino acids upstream (N-terminal) of VTLTCAFRY in the source proteins differ between the SAG and other arrestins (**Table 1**), we asked if this may influence the probability of antigen processing and presentation of the mature peptide VTLTCAFRY. To this end, we used neural network-based prediction of the the likelihood of the generation of N-terminally extended precursors peptides of VTLTCAFRY from full-length SAG, and □-arrestin-1 and -2 by the cellular antigen processing machinery (combined proteasomal and proteases) ^27,28^. This analysis revealed that whole-cell antigen processing is predicted to generate the 9-mer ligand and a N-terminally extended 11-mer for SAG, but not for other arrestins (**Fig 1D**). Of interest, the nearest N-terminal cleavage site “shared” between arrestins is predicted 5 amino acid positions before the 9-mer ligand, which would introduce N-terminal acidic residues (D and E) that are unfavourable for antigen processing ^35^. Collectively, these observations make it reasonable to assume that also the VTLTCAFRY peptide is derived specifically from SAG. Based on these results, we conclude that two 9-mer peptides derived from SAG are *bona fide* ligands of HLA-A29.

### HLA-A29 ligands of SAG are trimmed by ERAP1 and ERAP2

Because the antigen processing aminopeptidases ERAP1 and ERAP2 are genetically implicated in the pathogenesis of birdshot chorioretinitis ^7–9^, we next were interested to determine whether the two SAG-derived peptides could be generated through trimming N-terminal elongated peptide precursors by ERAP1 and ERAP2. We used recombinant ERAP1 and ERAP2 to *in vitro* digest VTLTCAFRY, TVLGILVSY and their respective 10-mer precursor peptides (N-terminal extended with the amino acid at that position in the retinal S-antigen amino acids sequence, **Table 2**). ERAP1 clearly processed the control peptides RGRFSGLLGR and SVLKSLPFTII, as well as the precursor peptide 10-mer YVTLTCAFRY to the 9-mer HLA-A29 ligand VTLTCAFRY after 60 minutes of incubation (**Fig. 3A,3B**). However, we observed no trimming of the 10-mer precursor RTVLGILVSY. No tryptic activity was detected in the ERAP preparations since the tryptic peptide SLPFTII peptide was not efficiently formed from SVLKSLPFTII. ERAP1 also trimmed the 9-mer HLA-A29 ligand VTLTCAFRY to the 8-mer TLTCAFRY. In contrast, ERAP2 left the precursor peptide 10-mer YVTLTCAFRY unaffected, but did show limited activity towards the precursor 10-mer RTVLGILVSY to yield the HLA-A29 ligand TVLGILVSY (**Fig. 3C**). These *in vitro* results show that SAG-derived ligands of HLA-A29 can potentially be generated from N-terminally trimming of precursor peptides by ERAP1 and ERAP2.

**Figure 3:**
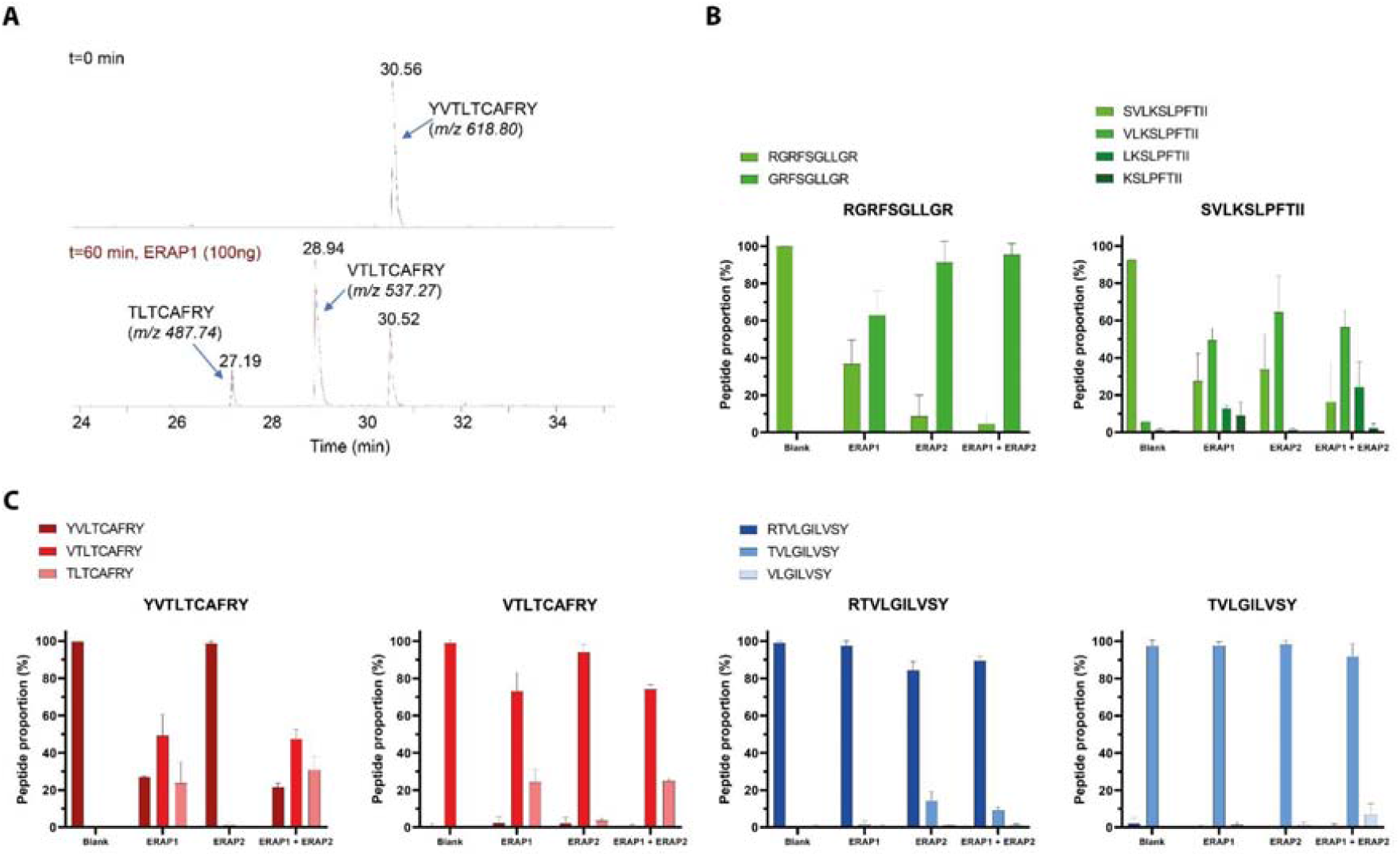
In vitro S-antigen peptide digestion by ERAP1 and ERAP2. (**A**) Representative mass spectrometry chromatogram showing ERAP1 digestion of S-antigen derived peptide YVTLTCAFRY into the respective 9- and 8-mer after the 60 min. incubation (**B**) Positive control RGRFSGLLGR, showing the proportional abundance digestion products (%) of the original peptide and measured digestion products as a result of ERAP1, ERAP2 or ERAP1 + ERAP2 digestion respectively. Control peptide SVLKSLPFTII to assess tryptic activity on lysine (K) at P4 to validate ERAP1/ERAP2 purity. **C**) Digestion of VTLTCAFRY and its respective 10-mer YVTLTCAFRY, TVLGILVSY and its respective 10-mer RTVLGILVSY. Peptide digestion assay data is shown in Supplemental Table 2.

### SAG-specific CD8+ T cells are not detectable in birdshot chorioretinitis patients

We next assessed whether the two identified peptides from the retinal S-antigen were detected by CD8+ T-cells from patients, HLA-A29-positive or HLA-A29-negative controls (n=34) using fluorescent-dextramerized recombinant HLA-A*29:02 molecules complexed with either VTLTCAFRY or TVLGILVSY. Comparing empty beads and beads coated with pan-class I antibody (W6/32) mixed with either one of the dextramers, revealed good performance for our assembled HLA-A29/VTLTCAFRY and HLA-A29/TVLGILVSY dextramers (**Fig. 4A**). Enumeration of dextramer-positive events revealed no difference between cases and HLA-A29-positive, and HLA-A29-negative controls (**Fig 4B**). Importantly, the frequency of detected events was in the same range as the negative control dextramer, indicating that no SAG-specific CD8+ T cells were detectable in peripheral blood.

**Figure 4.**
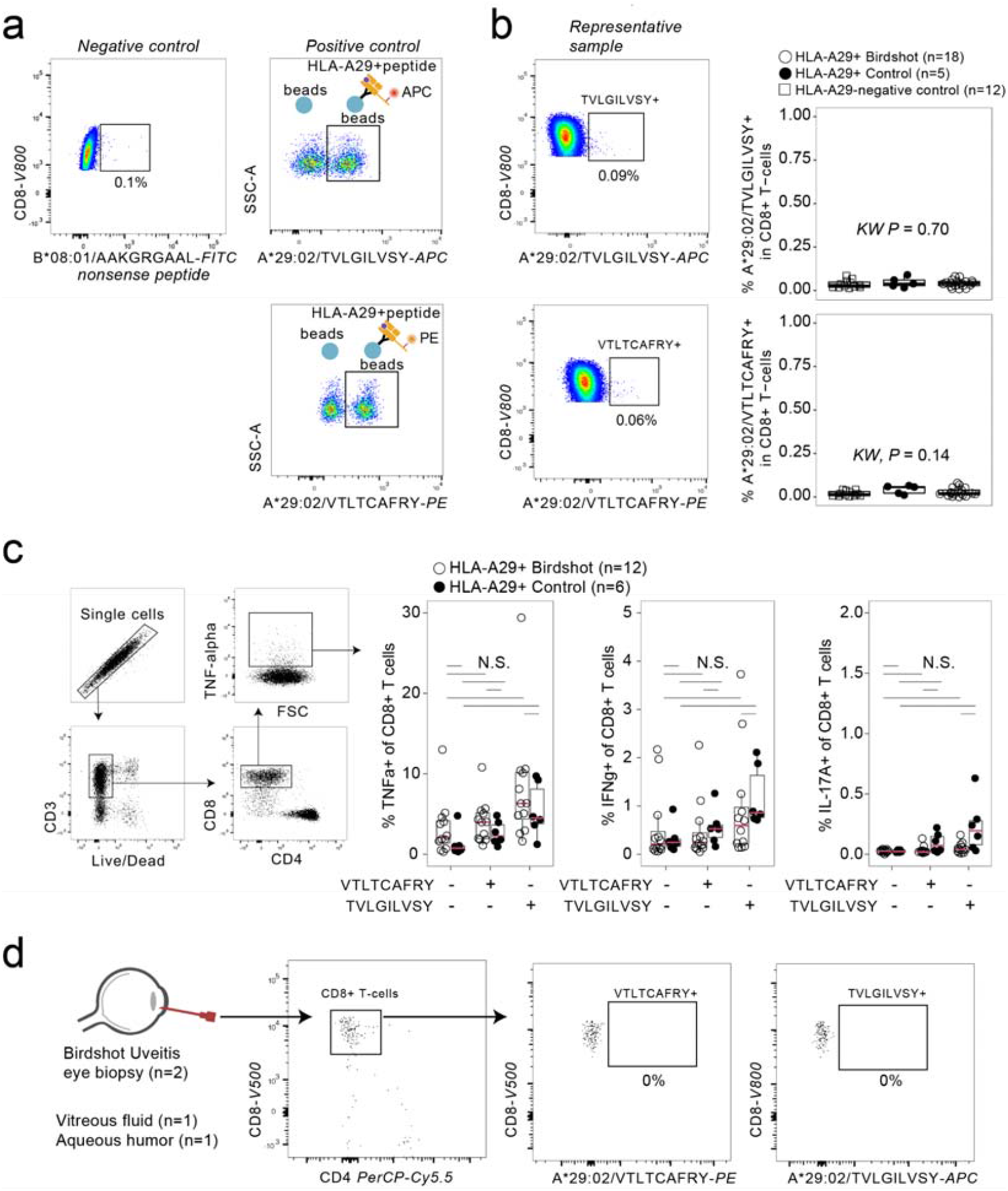
HLA-A29/SAG-specific CD8+ T cells are absent in peripheral blood or eye biopsies of birdshot chorioretinitis patients. **A)** Biplot of flow cytometry analysis using a positive control for HLA-A29-TVLGILVSY dextramer and HLA-A29-VTLTCAFRY dextramer by using beads coated with or without pan anti-HLA class I antibody (W6/32) and dextramers. As a negative control, we used a dextramer complexed with a nonsense peptide. **B**) Representative biplot for the percentage of the blood CD8+ T cells binding to each of the HLA-A29 dextramers. A scatterplot of the percentage of dextramer-positive CD8+T cells in peripheral blood mononuclear cell (PBMC) samples from HLA-A29+ birdshot chorioretinitis patients, HLA-A29+ healthy controls, and HLA-A29-negative healthy controls. *P* values are from a *kruskal wallis* test). **C**) Intracellular cytokine detection by flow cytometry of TNF-alpha, IFN-gamma, and IL-17-producing CD8+ T-cells after peptide stimulated PBMC cultures. The comparison of the percentage of cytokine producing CD8+ T-cells in unstimulated PBMC cultures, or stimulated with VTLTCAFRY, or TVLGILVSY, is shown. None of the comparisons were statistically significant by Dunn’s test with correction for multiple testing. NS = non-significant. **D**) Representative flow cytometry biplots of vitreous fluid of one birdshot chorioretinitis patient. Eye-infiltrating CD8+ T-cells were stained with the S-antigen peptide HLA-A29-TVLGILVSY dextramer and HLA-A29-VTLTCAFRY dextramer.

To ascertain the absence of CD8+ T cell activation by these SAG peptides, we assessed antigen engagement at the single cell level. To this end, we conducted peptide stimulations of PBMCs from 18 BCR patients and 6 HLA-A29-positive controls and monitored intracellular cytokine (TNF-alpha, interferon-gamma, and interleukin-17) production by CD8+ T-cells after 48 hours. Stimulation with synthetic VTLTCAFRY or TVLGILVSY did not induce a significant increase in the frequency of cytokine-producing CD8+ T-cells in cases and controls (**Fig. 4C**). Because we also detected VTLTCAFRY peptide with post-translational modifications in the immunopeptidome data, we also determined cytokine production by CD8+ T cells after 48 stimulation with VTLTCAFRY with phosphorylated Threonine (at amino acid position 2, position 4, or both position 2 and 4) or cysteinylated Cysteine (C-cys) at amino acid position 5 in VTLTCAFRY, but detected also no significant increase in the frequency of cytokine-producing CD8+ T cells in cases and controls using stimulation with these modified peptides (**Supplemental Figure 3**).

Finally, we had the opportunity to study fresh intraocular eye biopsies from two BCR patients. Although flow cytometry analysis revealed evident infiltration of CD8+ T cells, no SAG-peptide dextramer-positive T cells were detected in inflamed eyes of patients (**Fig 4D**). Collectively, we conclude that SAG peptides in complex with HLA-A29 are not recognized by CD8+ T-cells of BCR patients.

## Discussion

In this study, we investigated the candidate autoantigen SAG in BCR using patient-derived cell lines and identified two naturally presented peptides of SAG. We demonstrated that these peptides in complex with HLA-A29 are not recognized by circulating and eye-infiltrating CD8+ T cells of patients.

SAG has been postulated as the primary autoantigen ever since the discovery of HLA-A29 in cases with BCR over forty years ago ^18^. In 1982, Nussenblatt and coworkers established the extreme genetic association between *HLA-A*29* and BCR and concurrently reported that PBMC cultures of BCR patients showed enhanced proliferation upon stimulation with SAG compared to healthy controls ^18^. The nomination of SAG as candidate autoantigen for BCR was also based upon the observation that injecting rhesus monkeys with SAG or its derived 18-mer DTNLASSTIIKEGIDRTV (peptide “M”) induced experimental uveitis, characterised by focal retinal lesions and histopathological features with high resembles to BCR ^36,37^. However, *in vitro* SAG-stimulated T cell proliferation as well as SAG-induced experimental autoimmune uveitis is predominantly T helper cell-driven and mediated by MHC class II ^38–42^. Also, in humans, cellular and humoral *in vitro* hyperresponsiveness to SAG is not limited to BCR and commonly observed in patients with a wide variety of inflammatory eye conditions. Our study revealed that peptides from SAG presented by HLA-A29 are not immunogenic.

Using immunopeptidomics of newly-generated patients’ cell lines with low passage, we ensured preservation of canonical antigen processing for unbiased discovery of peptide antigens presented by HLA-A29. Although we consider this a significant improvement compared to more widely used long-established cell lines that suffer from genomic aberrations, also in the MHC-I pathway ^43,44^, ideally, we would have liked to determine if SAG ligands are also presented by photoreceptors in the eye under inflammatory conditions. Recently, it was shown that shortage in cellular tryptophan promotes tryptophan to phenylalanine substitution in proteins, which could alter the HLA antigenic peptide repertoire and activate T cells ^45^. HLA-A29 immunopeptidomics of inflamed retina is technically and ethically highly challenging considering the limited amount of tissue available for such analysis. However, even when considering the possibility of inflammation-induced tryptophan breakdown in the retina ^46–49^, SAG contains merely a single tryptophan in its amino acid sequence (W198), which after substitution does not influence predicted binding to HLA-A29 (NetCTLpan server after *in silico* W198F substitution, data not shown). We therefore consider it likely that our model captured all *in situ* SAG ligands of HLA-A29, also when presented by antigen presenting cells under inflammatory conditions.

Another limitation of our work is that we did not evaluate if CD8+ T cells recognize SAG-derived epitopes via other HLA class I alleles in linkage disequilibrium with HLA-A29 (e.g., HLA-B44, HLA-C16). However, in our reanalysis of previously reported pan-MHC-I immunopeptidome analysis of LCL1 ^21^ we detected only one other SAG-derived peptide (at Mascot Ion score >35): the 11-mer SEVATEVPFRL most likely derived from *HLA-B*44:03* (*HLAthena* binding score, MSI = 0.88) which is in linkage disequilibrium with *HLA-A*29:02* ^50^. Although the amino acid sequence of this 11-mer overlaps with peptides that are highly potent for induction of uveitis in a rat model of experimental autoimmune uveitis ^51,52^, this model is not mediated by CD8+ T cells ^53^. Therefore, we speculate that other putative HLA class I presented SAG epitopes will have negligible contribution to CD8+ T cell immunity in BCR. Given that PBMC cultures of BCR patients showed enhanced T cell proliferation upon stimulation with SAG^18,41^, our current view is that SAG functions as a “collateral” or “secondary” autoantigen involved in CD4+ T helper-mediated responses induced by epitope spreading subsequent to (yet unidentified) primary autoantigen-directed immunity via HLA-A29 and CD8+ T cells.

Peptide-MHC multimers represent a powerful technology for the detection of antigen-specific CD8+ T cells. However, the low frequency in peripheral blood and relatively low affinity T-cell receptors (TCR) of self-specific T-cells pose a significant challenge since the signal in peripheral blood may become obscured within the inherent background noise. To ascertain high quality detection and quantification, we used several strategies in line with current recommendations ^54^, such as an optimised staining protocol (stain with dextramers before staining for coreceptor CD8 ^55^), use of “dump” channels to eliminate nonspecific binding events^56^, FMO controls, and anti-multimer bead positive controls to ensure an accurate gating strategy. We also used dextramers which outperform conventional tetramers, because they carry a greater number of both peptide-HLA complexes and fluorochromes ^57^. Although the frequency of dextramer-positive events in this study were in the range of frequencies reported for blood CD8+ T-cells specific for viral or tumour peptides ^58–60^, we observed no differences to unaffected controls. Although we can formally not exclude HLA alloreactivity by CMV- and EBV-specific T-cells ^61–63^ known to cross-react specifically with HLA-A29 in HLA-A29-negative donors ^64^. Regardless, we demonstrated that the detected frequency in patients was similar to HLA-A29-positive controls, and indistinguishable from the negative control dextramer. Importantly, no dextramer-positive T cells were detected among the eye-infiltrating CD8+ T cells in patients. We also found no induction of antigen-specific cytokine production in CD8+ T cells after stimulation with these SAG peptides. This supports our conclusion that we did not detect SAG-specific CD8+ T cells in patients and that SAG is unlikely of relevance to the HLA-A29-mediated - and thus disease defining - pathogenic mechanisms.

Genome-wide genetic studies identified *HLA-A29, ERAP1*, and *ERAP2* genes as risk genes for BCR ^7,9^. This substantiates peptide processing by ERAP1 and ERAP2 in the endoplasmic reticulum and subsequent peptide presentation by HLA-A29 at the cell surface as a key disease mechanism for BU. Functional studies have revealed that ERAP2 promotes HLA-A29 presentation of peptides with a motif that is found in the amino acid sequence of melanocyte proteins ^1,21^. This is significant, because in *HLA-C*06-associated psoriasis* melanocytes trigger autoreactive CD8+ T cells via ERAP1 ^14,15^. These findings are significant because inflammation in BCR may originate in the melanocyte-rich ocular choroid and suggest that tissue resident autoantigens of the choroid may provide the targets for autoreactive CD8+ T cells ^1^. If melanocytes are a target tissue in BCR remains to be determined. Regardless, identification of autoantigens in upcoming studies may benefit from the rapid advancement in sequencing technologies, such as *single-cell TCR sequencing*, which will allow the identification of the full-length alpha and beta TCR chains of eye-infiltrating CD8+ T cells, which can be used for functional evaluation (i.e., cloning into cell lines) and screening of large peptide libraries to identify their cognate peptide antigen(s) ^15,65^.

In conclusion, we demonstrate that the longstanding autoantigen candidate SAG is not a CD8+ T cell autoantigen in BCR and that the disease mechanisms that involve antigen presentation via HLA-A29 involve other yet unidentified peptides.

## Data Availability

a dedicated DataverseNL dataverse upon publication.

https://dataverse.nl/

## Funding

JK is supported by a VENI award from the Netherlands Organization for Scientific Research (N.W.O. project number 016.186.006). WV is supported by UitZicht (project number 2018-1) and Stichting Lijf en Leven (project number 63). The funders had no role in the design, execution, interpretation, or writing of the study.

---

**Supplementary Figure 1.**
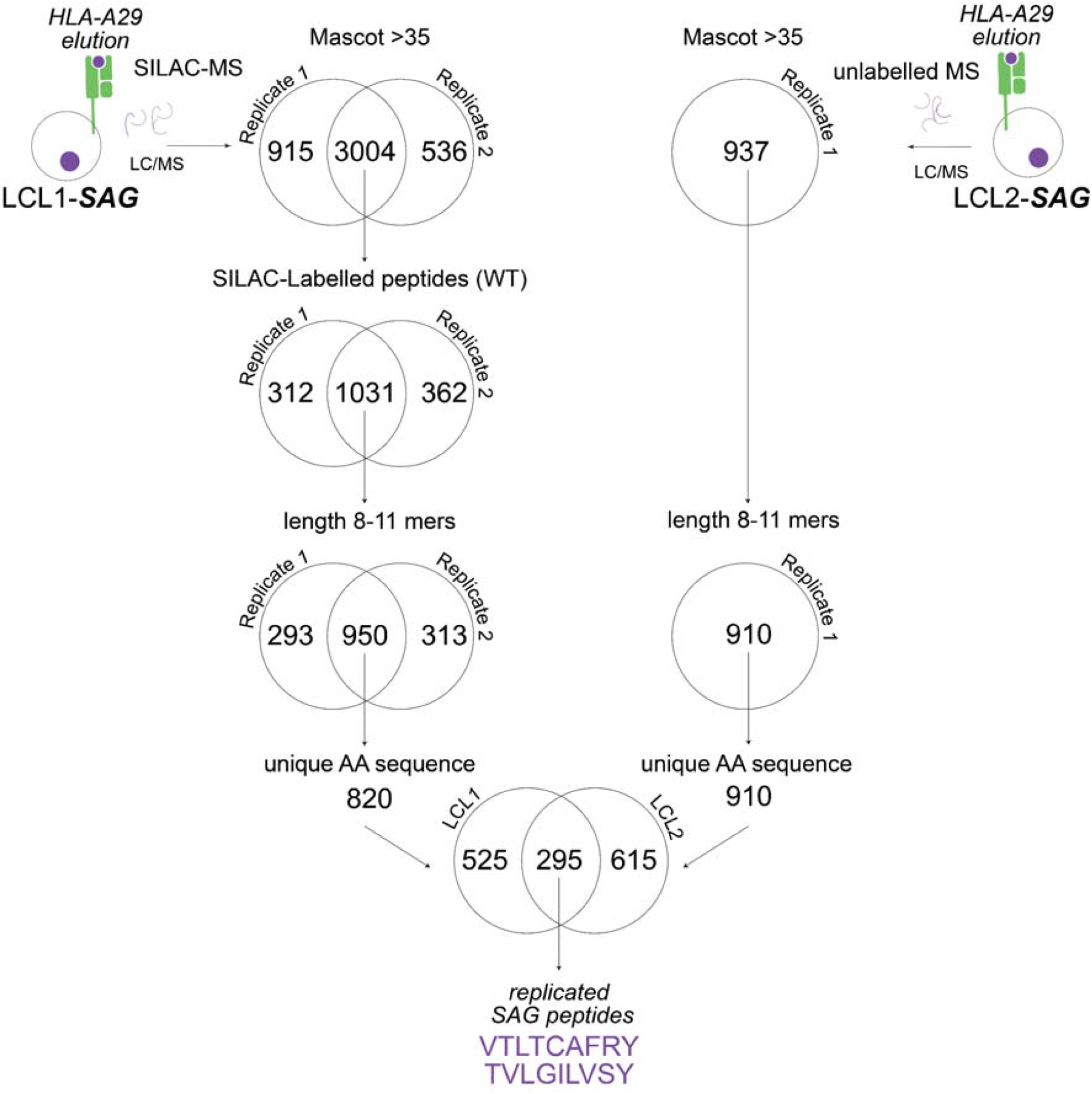
Venn diagram of filtering steps of the peptides identified by mass spectrometry in the HLA-A29 immunopeptidomes of LCL1 and LCL2.

**Supplementary Figure 2.**
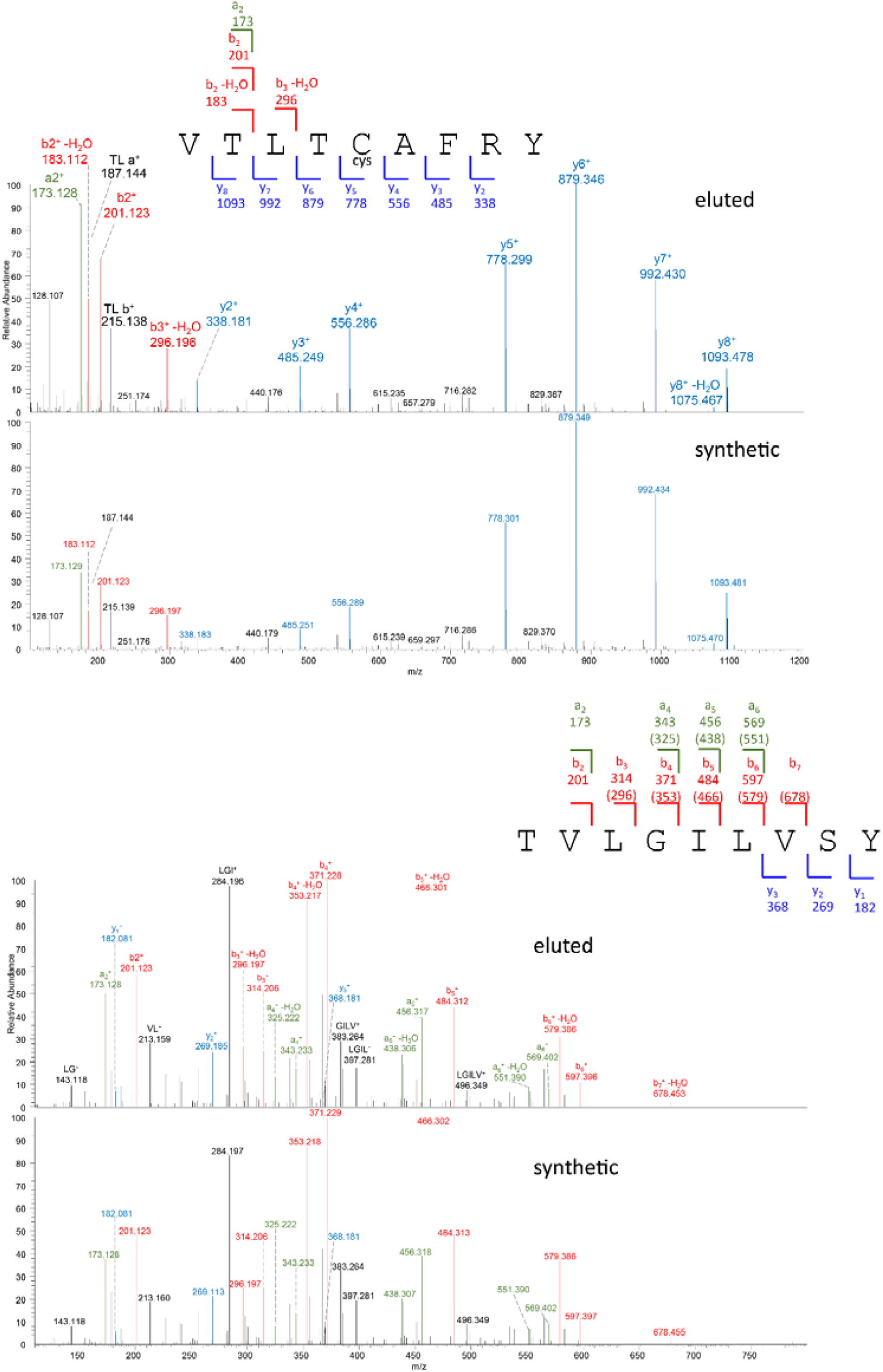
The MS/MS spectrum of the peptide fragment ions derived from the SAG peptides identified in elutions from HLA-A29 immunoprecipitations of the patient derived cell lines (“eluted”) and from recombinant peptides with identical amino acid sequence (“synthetic”). The relative abundance and mass/charge (m/z) of characteristic (a-,-b-, and y-) ions from the fragmentation of the peptides are highlighted.

**Supplementary Figure 3.**
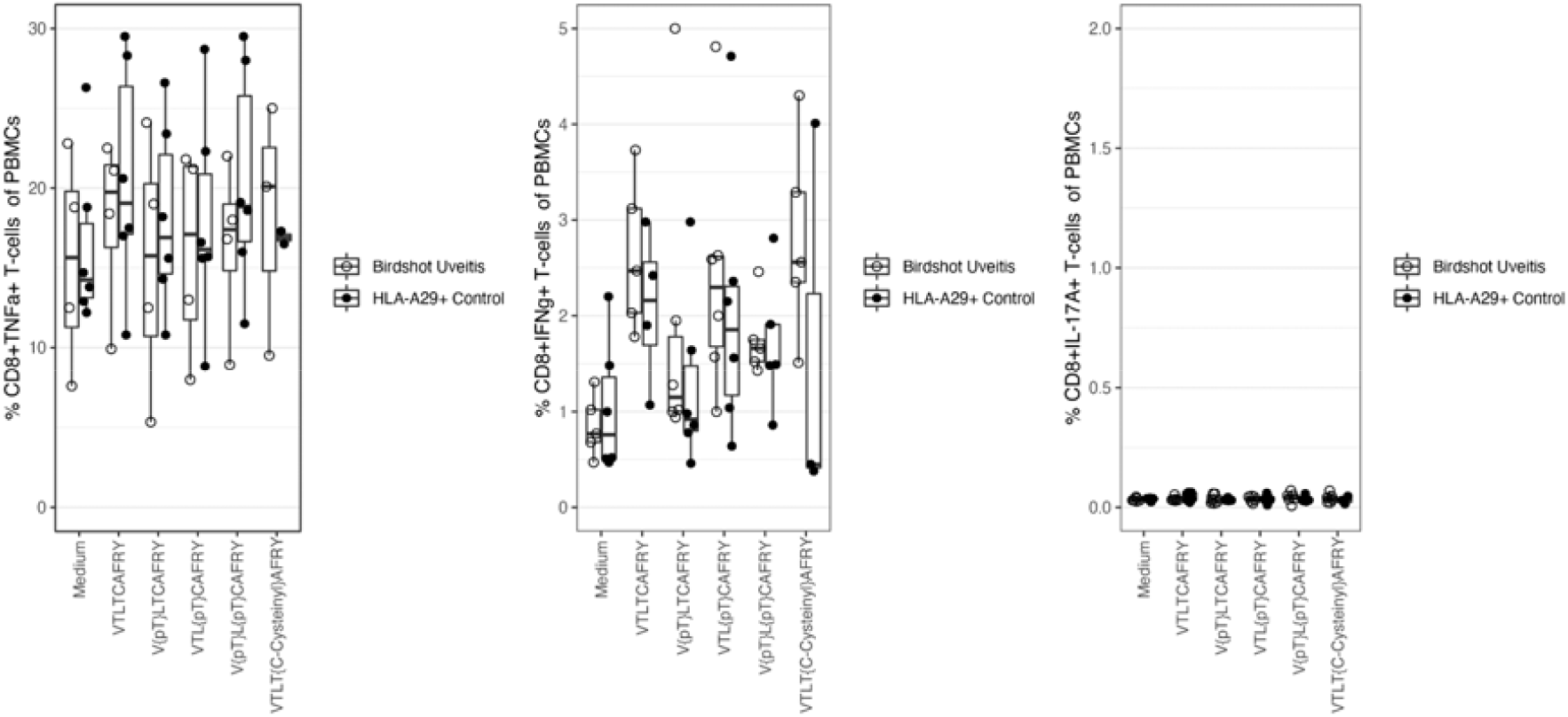
Intracellular cytokine detection by flow cytometry of TNF-alpha, IFN-gamma, and IL-17 producing CD8+ T-cells after S-antigen peptide stimulated cultures of PBMCs. The comparison of the percentage of cytokine producing CD8+ T-cells in unstimulated PBMC cultures, or stimulated with VTLTCAFRY, VTLTCAFRY with the phosphorylated T as position 2 (V{pT}LTCAFRY, VTLTCAFRY with the phosphorylated T as position 4 (VTL{pT}CAFRY, VTLTCAFRY with phosphorylated T as position 2 and 4 (V{pT}L{pT}CAFRY, and VTLTCAFRY with a cysteinyl to C at position 5 (VTLT{C-Cystenyl}AFRY is shown. None of the comparisons were statistically significant by Dunn’s test with correction for multiple testing.

## Supplemental Table 1-3

### Key Resources Table

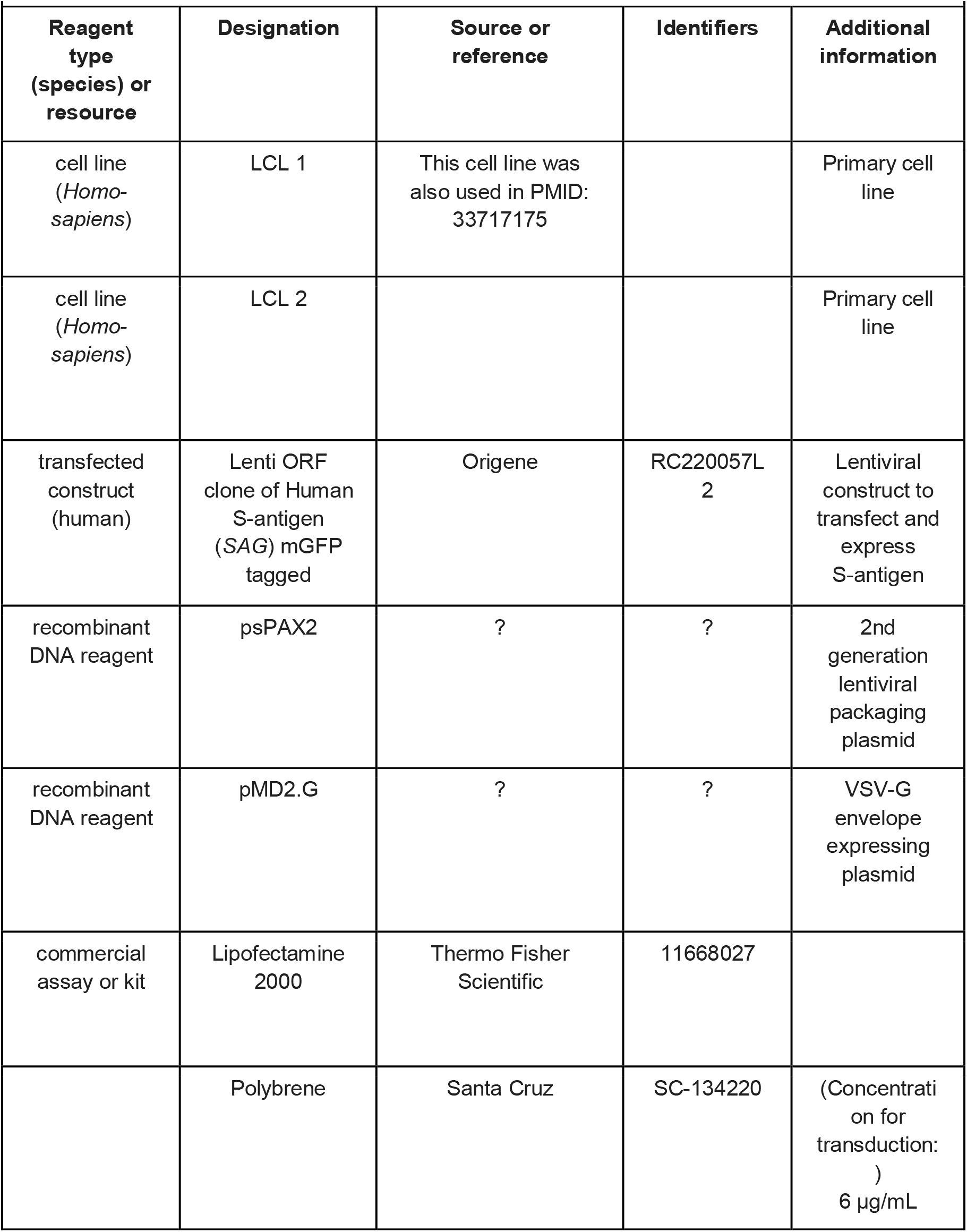
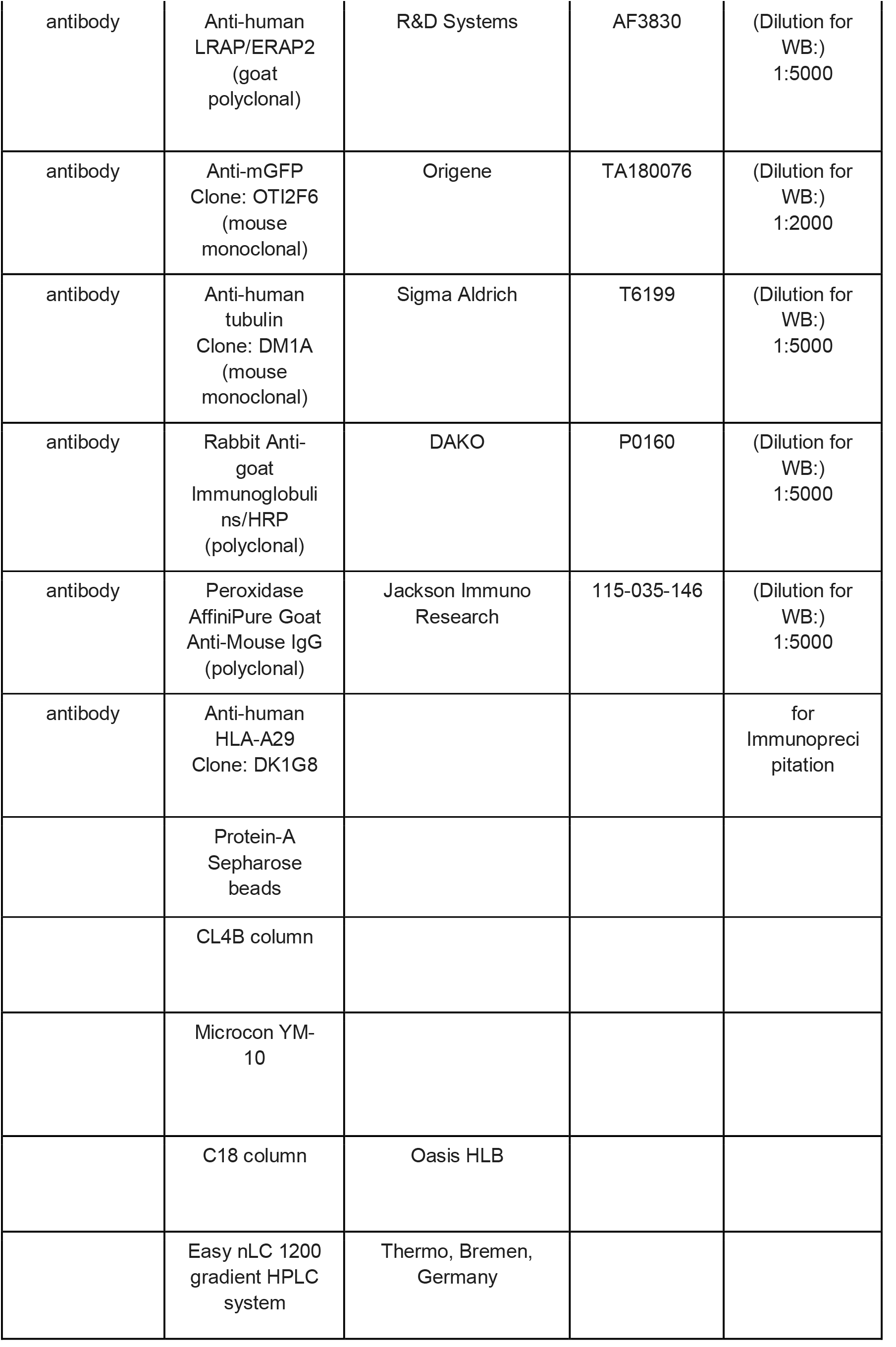
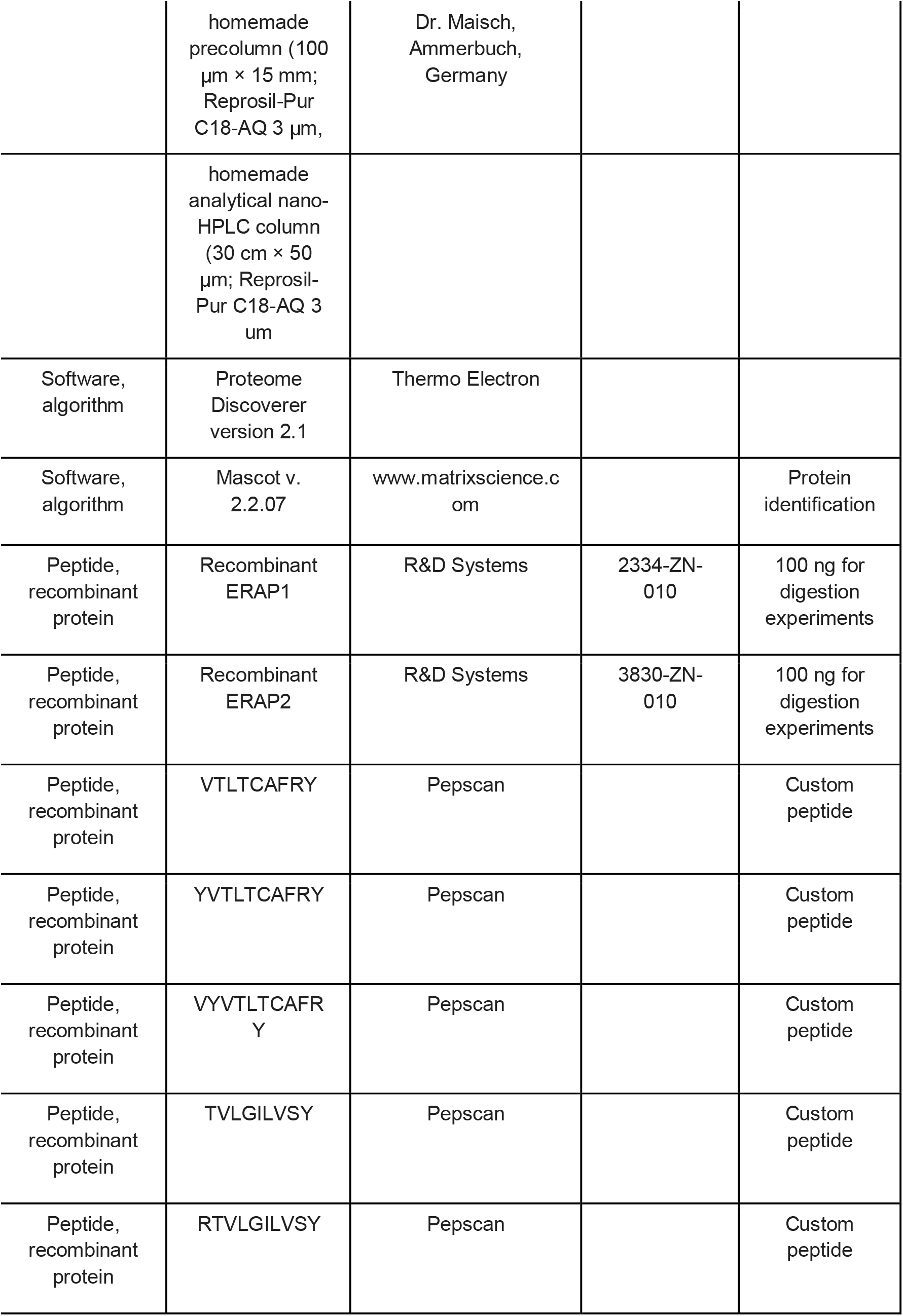
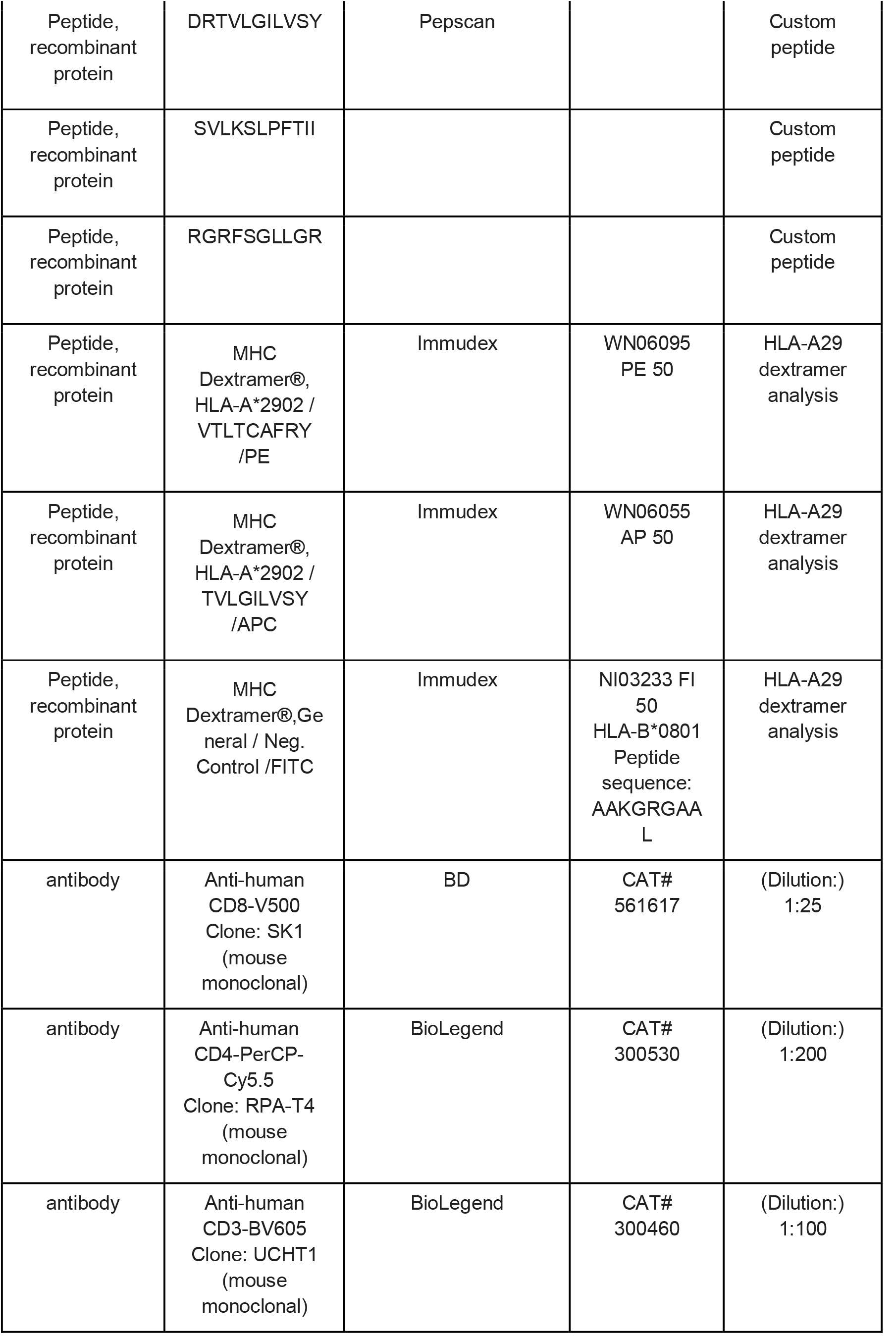
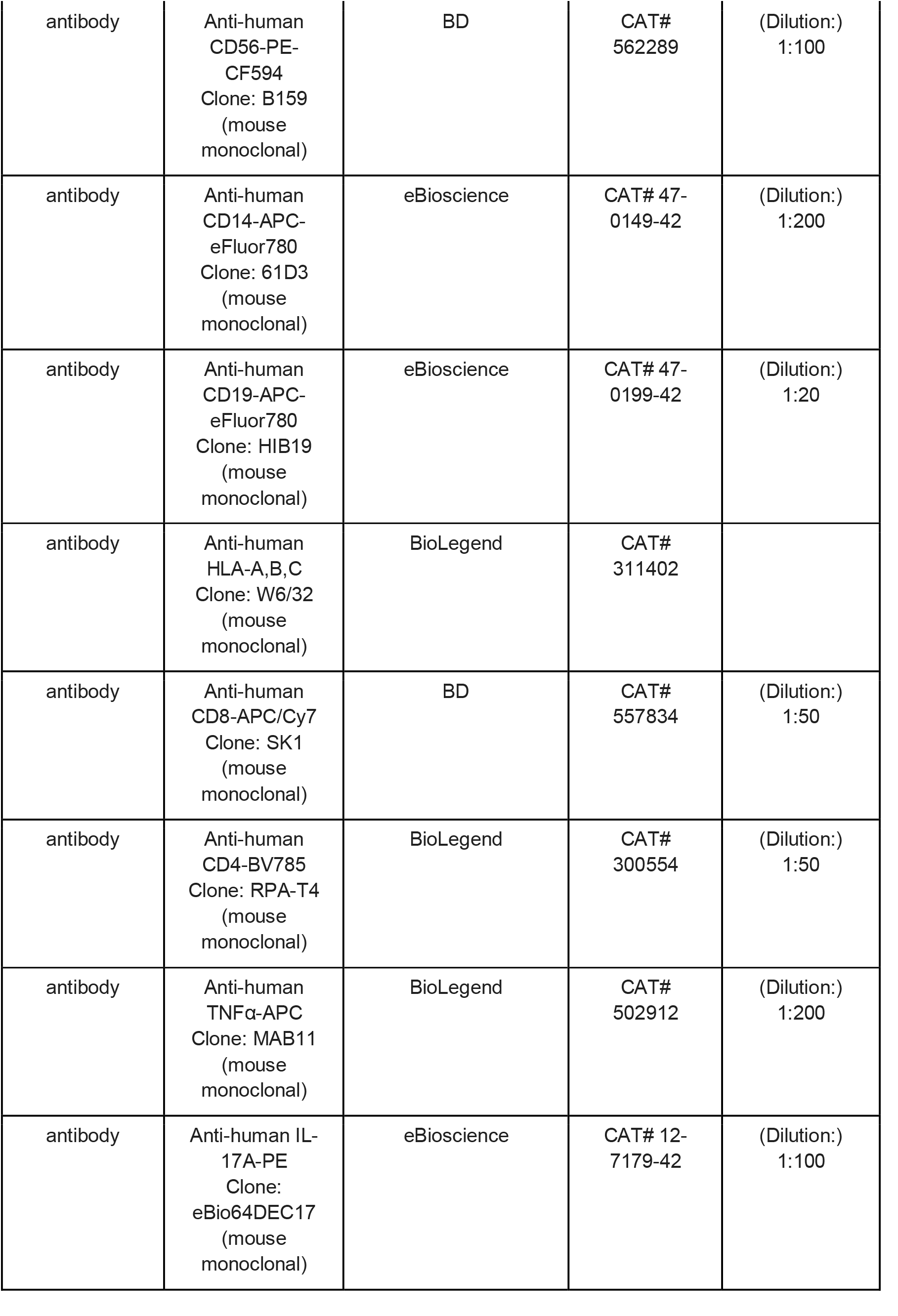
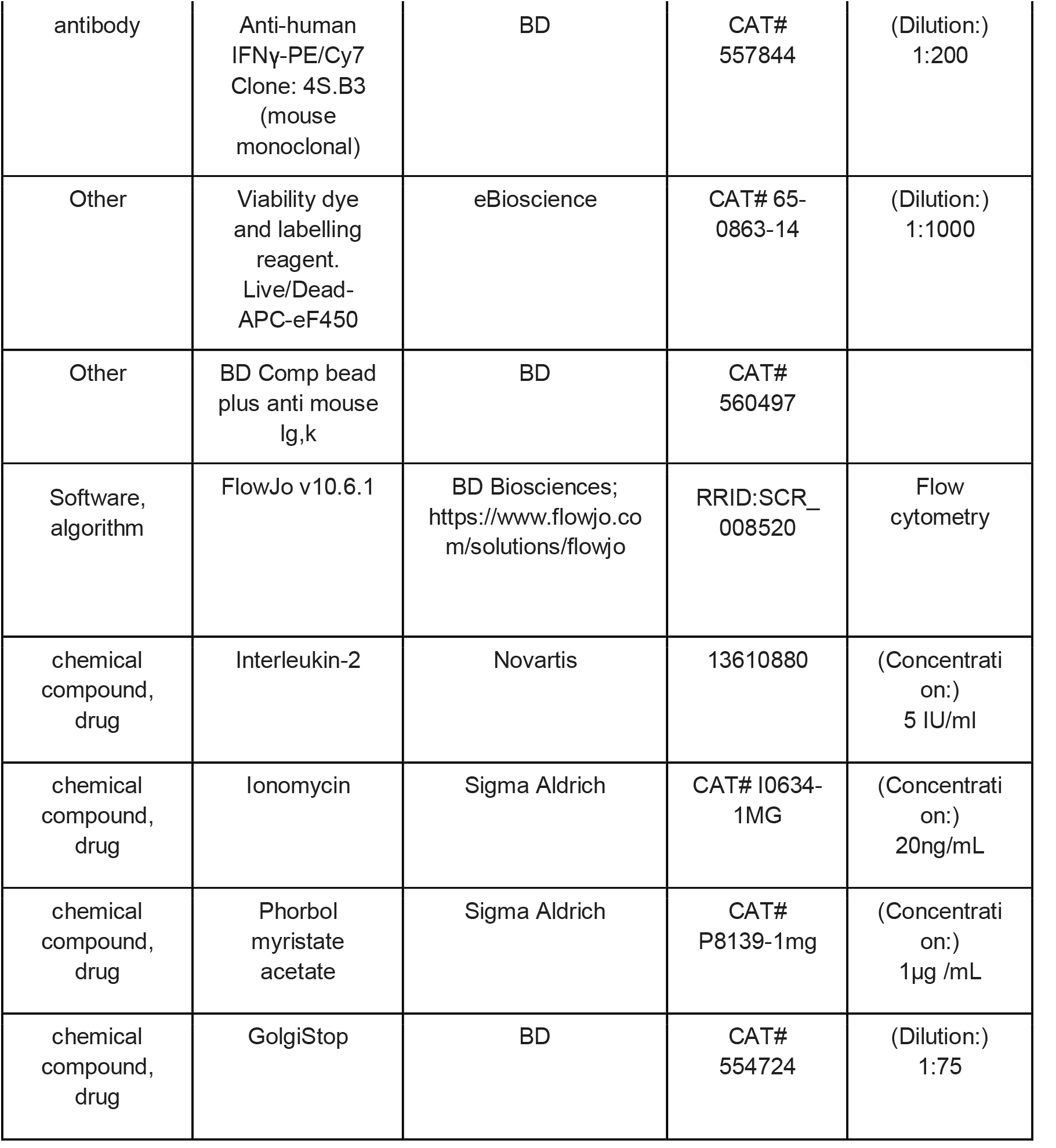

## Notes

### Competing Interest Statement

The authors have declared no competing interest.

### Author Declarations

The Medical Ethical committee of the University Medical Center Utrecht, the Netherlands, gave ethical approval for this work.

